# Improving Equity in Maternity Care Through Linguistically Accessible Parent Education Classes: A Proposal for London-Wide Mapping and Evaluation

**DOI:** 10.64898/2026.02.20.26346568

**Authors:** Heather Reeves, Miriam Bourke, Komal Khuti-Dullaart, Arezou Rezvani

## Abstract

**Background:** Women with limited or no English proficiency experience persistent barriers to accessing maternity care in the UK, contributing to well-documented inequalities in maternal and perinatal outcomes. NHS parent education classes are predominantly delivered in English, and provision of multilingual classes within individual maternity units is often limited and duplicative. Evidence to inform collaborative, cross-trust service models remains scarce.

**Objective:** To assess women’s access to, preferences for, and perceived relevance of NHS parent education classes, with particular focus on willingness to travel across London to attend classes delivered in a preferred language, in order to inform equitable and efficient service design.

**Methods:** A cross-sectional, multilingual survey was conducted as a quality improvement initiative across multiple London maternity networks. The survey was translated into 18 languages and captured sociodemographic characteristics, access to parent education, preferences for delivery format, timing, location, and language, and perceived relevance of content. Quantitative data were analysed descriptively and thematically.

**Results:** A total of 97 women participated in the survey (n=97), the majority of whom reported speaking at least one non-English language at home (79.4%, n=77). Regarding mode of delivery, 51.6% of women preferred in-person parent education classes (n=50), 15.5% preferred online delivery (n=15), and 32.9% reported no preference (n=32). Most participants reported access to a suitable device and reliable internet (85.6%, n=83) and confidence using online platforms (77.3%, n=75). In relation to timing and format, weekends were the most commonly preferred time for classes (40.2%, n=39), followed by weekdays during school hours (35.1%, n=34) and weekday evenings (24.7%, n=24). Nearly half of women preferred delivery across two 2-hour sessions (48.5%, n=47), while 30.9% reported no preference regarding session length or number (n=30); fewer preferred two 3-hour sessions (11.3%, n=11) or a single 4-hour session (8.3%, n=8). Regarding willingness to travel, 67.0% of participants reported they would attend parent education classes delivered outside their booking maternity unit (n=65). Overall, 68.0% were willing to travel up to 45 minutes for in-person classes (n=66), while 29.9% preferred not to travel (n=29). With respect to language of delivery, 40.2% of women preferred classes delivered in their native language (n=39), and a further 40.2% reported English with an interpreter as acceptable (n=39); fewer were comfortable relying on a partner or friend to translate (19.6%, n=19). Most participants perceived that delivery by a native-speaking health professional would improve trust and learning (75.3%, n=73), and an equal proportion expressed a preference for a female interpreter (75.3%, n=73).

**Conclusions:** Women with limited English proficiency demonstrate clear willingness to travel across maternity networks to access antenatal education in their preferred language. These findings support the development of collaborative, cross-trust models that standardise core antenatal content while centralising multilingual provision, reducing duplication and improving equity of access across London maternity services.

## Background

Like many other high-income countries, the United Kingdom has experienced sustained increases in migration over recent decades. By 2022, approximately 16% of the UK population was born outside the UK. In 2024, asylum seekers and refugees made up around 12% of immigrants to the UK (House of Commons Library, 2025). This demographic shift is reflected within maternity services: in 2023, 31.8% of all live births were to non-UK-born mothers in England and Wales. London remained the region with the greatest proportion of births, where 67.4% of live births were to parents where either one or both were born outside of the UK. Further, Around 47% of all foreign-born residents in the UK lived in London and the South East at the time of the 2021/22 Census (ONS, 2023). The latest Census report identified that 1.5% could not speak English well, and a 0.3% of the overall population in the UK could not speak English at all (ONS, 2022).

As a result, UK maternity services must respond to the specific needs of migrant women, particularly those who experience barriers to accessing care due to limited English proficiency. Evidence from the most recent MBRRACE-UK report (Felkner *et al.*, 2025) demonstrates that women from global majority communities experience significantly higher rates of adverse maternal and perinatal outcomes. Inadequate access to health information in a language women understand has been repeatedly identified as a contributing factor to these inequalities.

There is a well-established association between social deprivation and preexisting morbidities poorer maternal and neonatal outcomes. Language barriers intersect with these vulnerabilities, particularly for pregnant women requiring English interpretation. National policy reviews and inquiries, including the Three year delivery plan for maternity and neonatal services and HSIB investigations, have consistently identified communication failures as exacerbating factors in adverse maternity outcomes (NHS England, 2023; HSIB, 2020). Essential Action 7 of the Ockenden Review specifies that women must be informed of their birth options in a way they can understand, explicitly recognising communication and language as critical determinants of informed choice. Further guidance from the Royal College of Obstetricians and Gynaecologists (RCOG, 2025) reinforces the requirement for appropriate language support within maternity services.

Over the past decade, maternal mortality has disproportionately affected women from Black and minority ethnic backgrounds. Compared to White women, women of Asian ethnicity are 1.3 times more likely and women of Black ethnicity 2.3 times more likely to die within six weeks of childbirth. The 2021 and 2023 Confidential Enquiries into Maternal Deaths found that 9.34% of women who died did not speak English. Limited access to understandable health information including medication adherence, appointment attendance, and perinatal screening significantly impacts outcomes (Felker *et al*., 2025).

Language barriers are consistently linked to poorer maternal and neonatal outcomes (Turin *et al*., 2021). Kenyan *et al.* (2024), review into 25 maternal deaths among recent migrant women with no or limited English, highlighted limited research on improving outcomes for this group and recommended raising awareness among this group of how to navigate maternity services, and how to both recognise and access pregnancy related emergencies. This crucial information is often missed when antenatal education is only provided in English. Providing health information in a comprehensible way is central to equitable maternity care, helping women understand antenatal pathways, screenings, and services, advocate for interpreters, and seek second opinions empowering them and reducing disparities in access and outcomes.

Family dynamics and cultural expectations significantly influence engagement with maternity services. Intergenerational beliefs and the views of partners or in-laws can downplay the importance of antenatal care (Goodwin et al., 2017). Migrant women often navigate parallel belief systems, combining traditional practices with Western healthcare (Woollett et al., 1995).Nagesh *et al*. (2024) emphasises the importance of extending education beyond expectant parents to include female in-laws, noting that tensions between traditional practices and evidence-based maternity care can create conflict for migrant women, particularly when living close to extended family members. These findings highlight the complexity of delivering maternity education that is both culturally sensitive and evidence based.

Effective interventions include tailored education for families, with medical terminology in native languages. The Health Literacy Project in Tower Hamlets (Inspire the Mind, 2025) offered Bangla-speaking women classes on pregnancy-related English, anatomy, and maternity care, using ESOL teachers, healthcare professionals, community researchers, visual aids, and role-play. The program was well received (unpublished findings) and is expanding to additional languages.

In parallel, emerging evidence highlights the increasing role of digital misinformation in shaping expectant parents’ knowledge and decision-making. A growing proportion of online maternity-related content, particularly on social media platforms contains misinformation or commercially motivated messaging, including the promotion of non-evidence-based products. Despite this, the digital health information landscape remains largely unregulated. This context underscores the responsibility of the NHS to provide free, evidence-based, and linguistically accessible maternity information, free from commercial influence to all families regardless of language proficiency.

Practically speaking, it is not financially viable to provide parent education classes in a variety of languages. However, service provisions that work on collaboration rather than duplication may be able to extend the provisions available to this vulnerable group of women. An example is the maternity Unit within zone 1 London that this project was registered under, there are 49 recorded languages amongst our pregnant users. Further, maternity units do not share resources and instead duplicate services. The research proposes a service provision that forms partnerships. The exploratory survey is to review the feasibility for maternity units within a reasonable commutable distance to agree standardisation of the health information that all women need to have, and then to agree specialisation language hubs and signpost women to the antenatal class.

## Overall aim/ Primary Objective

To evaluate access to, preferences for, and perceived relevance of NHS parent education classes among women with limited or no English proficiency, in order to inform equitable service design.

## Secondary objectives

To identify barriers to attendance and opportunities for service redesign that may improve equity of access to parent education classes.

## Aim

1. What are women’s preferences for the mode of delivery (online vs in-person) of NHS parent education classes, and how do digital access and confidence with online platforms influence these preferences?
2. What are women’s preferred timing (time of day, day of week), duration, and number of parent education sessions?
3. To what extent are women willing to attend parent education classes delivered outside their booking maternity unit or at central London locations, and how far are they willing to travel?
4. What are women’s preferences regarding the language of delivery (native language, English with interpreter, or partner-supported translation)?
5. Do women perceive that delivery by a native-speaking midwife or health professional improves trust and learning, and what is the preference for a female interpreter?

## Method

This project was registered as a quality improvement initiative and aligns with national policy priorities. Education leads across London and the maternity workforce at the host Trust were invited to participate. A working group including consultant and lead midwives, perinatal quality and mortality leads, antenatal ward and clinic leads, and the lead Maternity & Neonatal Voice Partner (MNVP) developed the survey questions. MNVPs, which embed service user voices in maternity and neonatal care, actively reframed questions based on women’s feedback.

The Iolanthe Midwifery Trust, supported by a generous Dora Opoku award, funded this public involvement project. Each participant received a £10 Amazon e-voucher. Following NIHR (2024) guidance, fair reimbursement ensures contributions are valued and a diverse range of voices is heard. The reimbursement aligns with recommended amounts and is intended to support inclusive participation rather than act as an incentive. Participation in the survey was emphasised as voluntary, highlighted that by completing the survey, would not affect the care women received, and that implied consent would be taken on completion of the survey. No identifiable information was collected. It was disclaimed that anonymised responses may use for publications or to improve maternity services.

Microsoft forms was used to generate an English version of the survey, comprising of 49 questions, (divided into qualitative and thematic analysis) and then, with the use of Google Translate, the survey was translated into the 18 top languages within maternity units. six of the surveys were cross referenced for language accuracy. The questions posed were intentionally all structured to accommodate those with limited English or low literacy levels and were set for the national reading level of the UK, which is in line with the latest PIAAC report (Department of Education, 2024) and promotes inclusivity.

This analysis uses cross-sectional survey data collected from women receiving maternity care. The survey captured sociodemographic characteristics, access to and attendance at parent education classes, perceived relevance of class content, and free-text feedback on service improvement. Survey data were analysed using descriptive, comparative, and multivariable methods. Participant characteristics were summarised using counts (n=) and percentages (%) for categorical variables and medians with 25-75th interquartile ranges (IQRs) for continuous variables. Access to parent education classes was assessed by describing whether classes were offered, gestational age at which they were offered, and attendance, both overall and stratified by key equity-related characteristics, including language spoken at home, migration status, education level, parity, and hospital site.

Perceived relevance of parent education content was summarised descriptively, with subgroup comparisons conducted where appropriate. Free-text responses were analysed thematically to identify barriers, facilitators, and suggested improvements to services, supported by illustrative quotations. The extent of missing data was assessed and reported for all key variables; analyses were conducted using complete-case analysis without imputation, and patterns of missingness were considered in interpretation. All analyses were conducted using Stata version 18.5, with a two-sided significance level of 0.05

## Results

### Participant characteristics

A total of 97 women completed the survey (Table 1). Participants were recruited across multiple London maternity networks, with the largest proportions from North Central London (35.8%) and North West London (27.4%). Consistent with the study focus, most participants spoke at least one non-English language at home (79.4%). Bengali (28.9%), Spanish (15.5%), Arabic (15.5%), and Urdu (10.3%) were the most frequently reported languages, participants could select multiple languages. While 20 women (20.6%) reported speaking English at home, this was typically in addition to another language, rather than as the sole household language (figure 1). The sample was ethnically diverse, with the majority identifying as Asian (23.7%) or Mixed/Other ethnicity (35.1%), and smaller proportions identifying as Black (5.2%) or White (1.0%). Over half reported temporary or insecure migration status (50.5%), while 17.5% identified as British or UK citizens. Half of participants had university-level education or higher (50.5%), while 13.4% reported primary education or no formal education. Most women were married (75.3%) or living with a partner or family member (13.4%), with a small proportion reporting single status (1.0%). Approximately half were nulliparous (49.5%), and responses were evenly distributed across pregnancy stages, with 38.1% postnatal at the time of survey completion. At the time of survey completion, women were at varying stages of pregnancy or early motherhood: 23.7% were ≤30 weeks pregnant, 37.1% were ≥31 weeks pregnant, and 38.1% had given birth within the previous two years.

**Figure 1:**
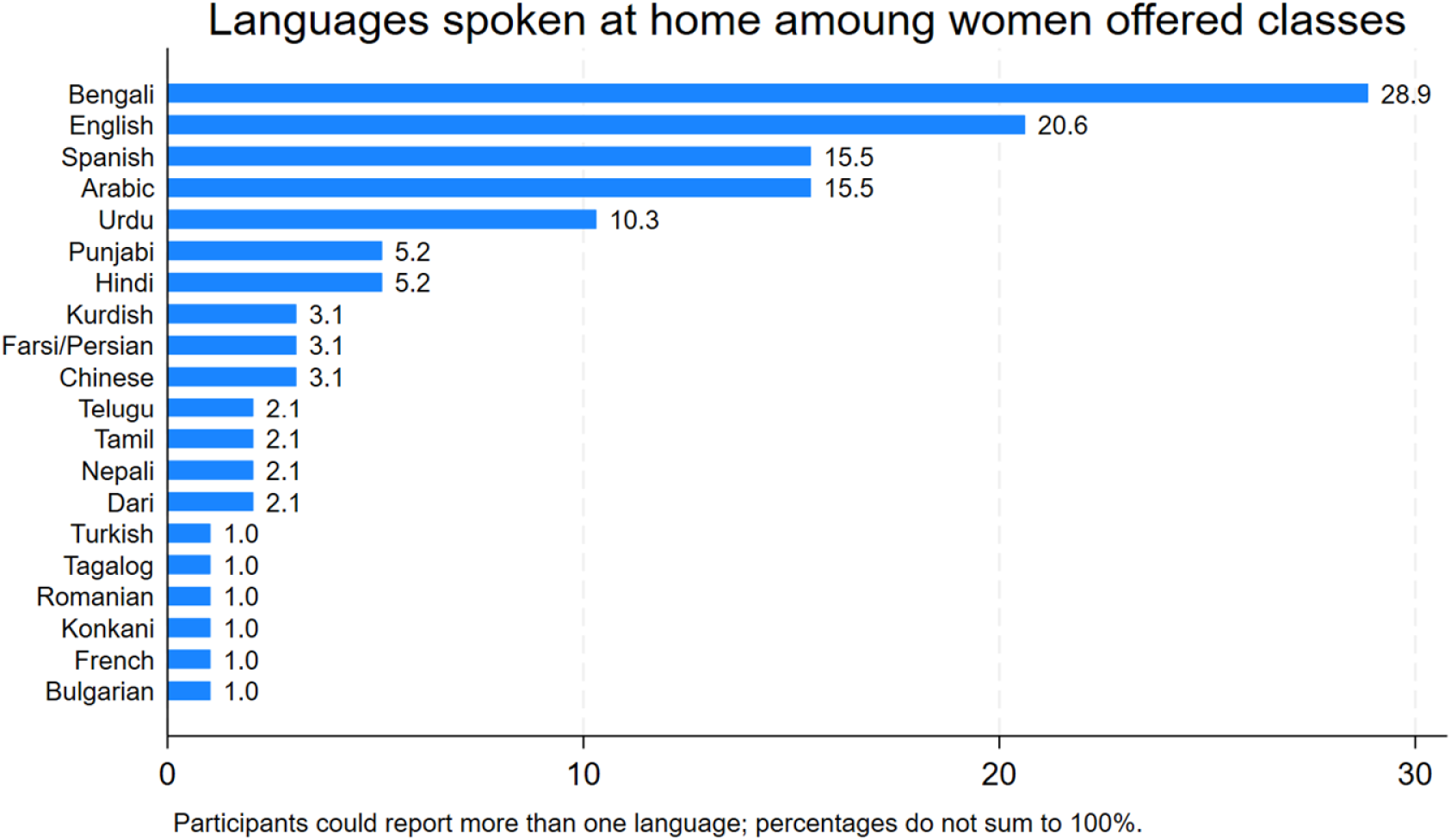
**Languages spoken at home among survey participants; Participants could report more than one language; percentages therefore do not sum to 100%.**

**Table 1.** Participant sociodemographic and pregnancy characteristics (Q2-4, Q37-42)

| Characteristic | Total (N=97)<br>n (%) |
| --- | --- |
| Hospital site (Q2) |  |
| North East London LMNS | 18 (18.9) |
| North Central London LMNS | 34 (35.8) |
| North West London LMNS | 26 (27.4) |
| South East London LMNS | 3 (3.2) |
| South West London LMNS | 8 (8.4) |
| Outside London LMNS | 6 (6.3) |
| **Language spoken at home\* (Q3)** |  |
| Bengali* | 28 (28.9) |
| English with another language* | 20 (20.6) |
| Arabic* | 15 (15.5) |
| Spanish* | 15 (15.5) |
| Urdu* | 10 (10.3) |
| Punjabi* | 5 (5.2) |
| Hindi* | 5 (5.2) |
| Chinese* | 3 (3.1) |
| Kurdish* | 3 (3.1) |
| Farsi* | 3 (3.1) |
| Dari* | 2 (2.1) |
| Tamil* | 2 (2.1) |
| Telugu* | 2 (2.1) |
| Nepali* | 2 (2.1) |
| Turkish* | 1 (1.0) |
| Tagalog* | 1 (1.0) |
| Romanian* | 1 (1.0) |
| Bulgarian* | 1 (1.0) |
| Konkani* | 1 (1.0) |
| Hungarian* | 1 (1.0) |
| French* | 1 (1.0) |
| **Ethnicity (Q37)** |  |
| White | 1 (1.0) |
| Black | 5 (5.2) |
| Asian | 23 (23.7) |
| Mixed/Other | 34 (35.1) |
| **Migration status (Q42)** |  |
| British/UK citizen | 17 (17.5) |
| Secure status/long-term resident | 15 (15.5) |
| Temporary/insecure status | 49 (50.5) |
| **Highest Education level (Q40)** |  |
| No formal education | 2 (2.1) |
| Primary School (school at age 11) | 11 (11.3) |
| Secondary school/ Higher education | 27 (27.8) |
| University degree or higher | 49 (50.5) |
| **Marital Status** |  |
| Married | 73 (75.3) |
| Living with partner/family | 13 (13.4) |
| Single | 3 (1.0) |
| **Parity (Q38)** |  |
| Nulliparous | 48 (49.5) |
| Multiparous ($\geq 1$) | 41 (42.3) |
| **Pregnancy gestation at survey (Q4)** |  |
| $\leq 30$ weeks pregnant | 23 (23.7) |
| $\geq 31$ weeks pregnant | 36 (37.1) |
| Postnatal ($\leq 2$ years) | 37 (38.1) |

\* Participants could report more than one language.
Percentages are calculated using available data. Completion of some sociodemographic questions was optional; missingness ranged from 8% to 53% across variables.

#### Access to NHS parent education classes

Overall, 78 women (80.4%) reported being offered NHS parent education classes (Table 2). Among those offered classes, the median gestational age at which classes were offered was 28 weeks (IQR 20–31 weeks). Most women perceived the timing of the offer as “just right” (85.6%), although 9 women (2.8%) felt classes were offered too late. When asked about preferred timing, women expressed a desire for earlier access to parent education. Over two-thirds indicated a preference for classes to start before 32 weeks’ gestation, with 25.8% preferring access as early as possible and a further 55.6% preferring initiation between 20 and 32 weeks. Only 12.4% preferred classes to start at or after 33 weeks, and 6.2% reported no preference.

**Table 2:** Access to NHS parent education classes, timing, and language provision (Q5-9)

| **Outcome** | **n (%)** |
| --- | --- |
| Offered NHS parent education classes (Yes) (Q5) | 78 (80.4) |
| Gestational age when classes were offered (weeks), median (IQR) (Q6) | 28 (20-31) |
| Timing perceived as (Q7) |  |
| Just right | 83 (85.6) |
| Too early | 1 (1.0) |
| Too late | 9 (2.8) |
| Preferred gestational age to start classes (Q8) |  |
| As early as possible | 25 (25.8) |
| 20-25 weeks | 16 (16.5) |
| 26-28 weeks | 14 (14.4) |
| 29-32 weeks | 24 (24.7) |
| 33-36 weeks | 9 (9.28) |
| After 37 weeks | 3 (3.1) |
| No preference | 6 (6.2) |
| Language(s) classes offered in (Q10) |  |
| English with interpreter | 44 (45.4) |
| English with friend/partner translating | 4 (4.1) |
| Urdu | 2 (2.1) |
| Bengali | 14 (14.4) |
| Spanish | 10 (10.3) |
| Arabic | 2 (2.1) |
| Not offered | 18 (18.6) |
| Midwife explained importance / recommended attendance (Yes) (Q9) | 83 (85.6) |
| Willing to attend classes outside own maternity unit (Q13) | 65 (67.0) |

Regarding language provision, fewer than half of women reported that classes were offered in English with an interpreter (45.4%), while smaller proportions reported classes offered directly in Bengali (14.4%), Spanish (10.3%), Arabic (2.1%), or Urdu (2.1%). Nearly one in five women (18.6%) reported that parent education classes were not offered to them at all.

The majority of women (85.6%) reported that a midwife had explained the importance of parent education or recommended attendance. Two-thirds of participants (67.0%) indicated they would be willing to attend classes delivered outside their booking maternity unit.

### Preferences for delivery, timing, and format

Preferences for class delivery and format are summarised in Table 3. Over half of women preferred in-person classes (51.6%), while 15.5% preferred online delivery and 32.9% had no preference. Most participants reported having access to a suitable device and reliable internet (85.6%) and felt confident using online platforms (77.3%). Preferred timing varied, with weekends being the most commonly preferred option (40.2%), followed by weekdays during school hours (35.1%) and weekday evenings (24.7%). Nearly half of participants preferred delivery across two 2-hour sessions (48.5%), while almost one-third reported no preference regarding session length or number (30.9%). Fewer participants favoured two 3-hour sessions (11.3%) or a single 4-hour session (8.3%). Willingness to travel for in-person classes is shown in Figure 2. While 68.0% of women were willing to travel up to 45 minutes, nearly one-third (29.9%) reported that they would prefer not to travel for in-person classes.

**Figure 2:**
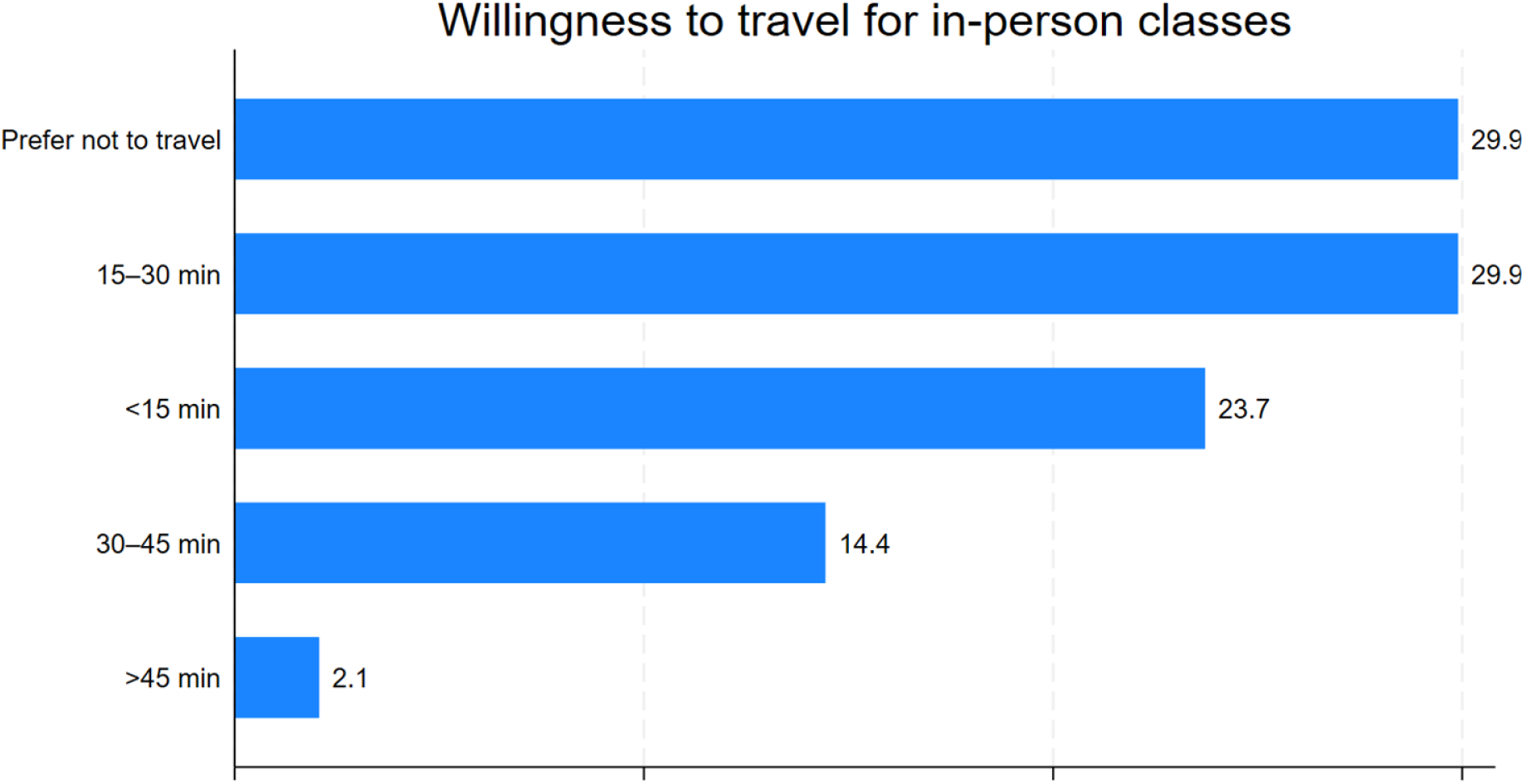
**Willingness to travel to attend in-person NHS parent education classes**

**Table 3:** Preferences for delivery, format, language, and facilitation of NHS parent education classes.

| Domain | Measure | n (%) |
| --- | --- | --- |
| Class mode of delivery (Q11) | Prefer online | 15 (15.5) |
|  | Prefer in-person | 50 (51.6) |
|  | No preference | 32 (32.9) |
| Digital access (Q21) | Has device + reliable internet | 83 (85.6) |
|  | No reliable device/internet | 14 (14.4) |
| Confidence with online platforms (Q22) | Confident using online platforms | 75 (77.3) |
|  | Not confident | 22 (22.7) |
| Preferred timing of classes (Q18) | Weekdays (school hours) | 34 (35.1) |
|  | Weekday evenings | 24 (24.7) |
|  | Weekends | 39 (40.2) |
| Preferred format/duration (Q19) | 2-hour sessions × 2 | 47 (48.5) |
|  | 3-hour sessions × 2 | 11 (11.3) |
|  | Single 4-hour session | 8 (8.3) |
|  | No preference | 30 (30.9) |
| **Language of delivery (Q12)** | Native language preferred | 39 (40.2) |
|  | English with interpreter acceptable | 39 (40.2) |
|  | English with partner/friend translating acceptable | 19 (19.6) |
| **Willingness to travel (Q14)** | <15 minutes | 23 (23.7) |
|  | 15-30 minutes | 29 (29.9) |
|  | 30-45 minutes | 14 (14.4) |
|  | >45 minutes | 2 (2.06) |
|  | Prefer not to travel | 29 (29.9) |
| **Trust and facilitation (Q15/16)** | Native-speaking professional improves trust (Yes) | 73 (75.3) |
|  | Preference for female interpreter (Yes) | 73 (75.3) |

#### Language of delivery and trust

Preferences for language of delivery are presented in Table 3 and Figure 3. 40% of women preferred classes delivered in their native language (40.2%), while a further 40.2% reported that English with an interpreter would be acceptable. Fewer women (19.6%) were comfortable relying on a partner or friend to translate. Most participants perceived that delivery by a native-speaking midwife or health professional would improve trust and learning (75.3%), and an equal proportion expressed a preference for a female interpreter (75.3%) (Figure 4).

**Figure 3:**
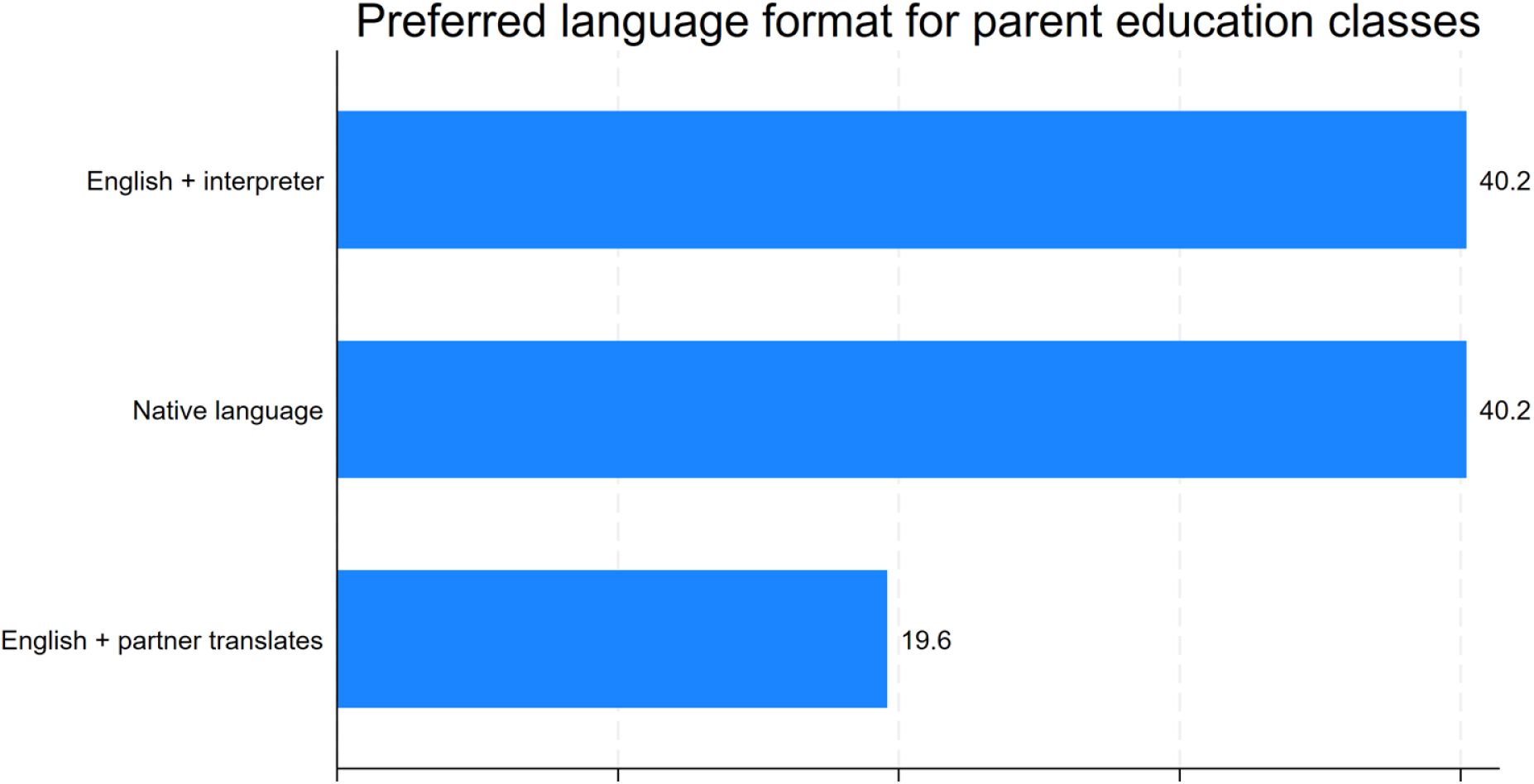
**Preferred language format for NHS parent education classes**

**Figure 4:**
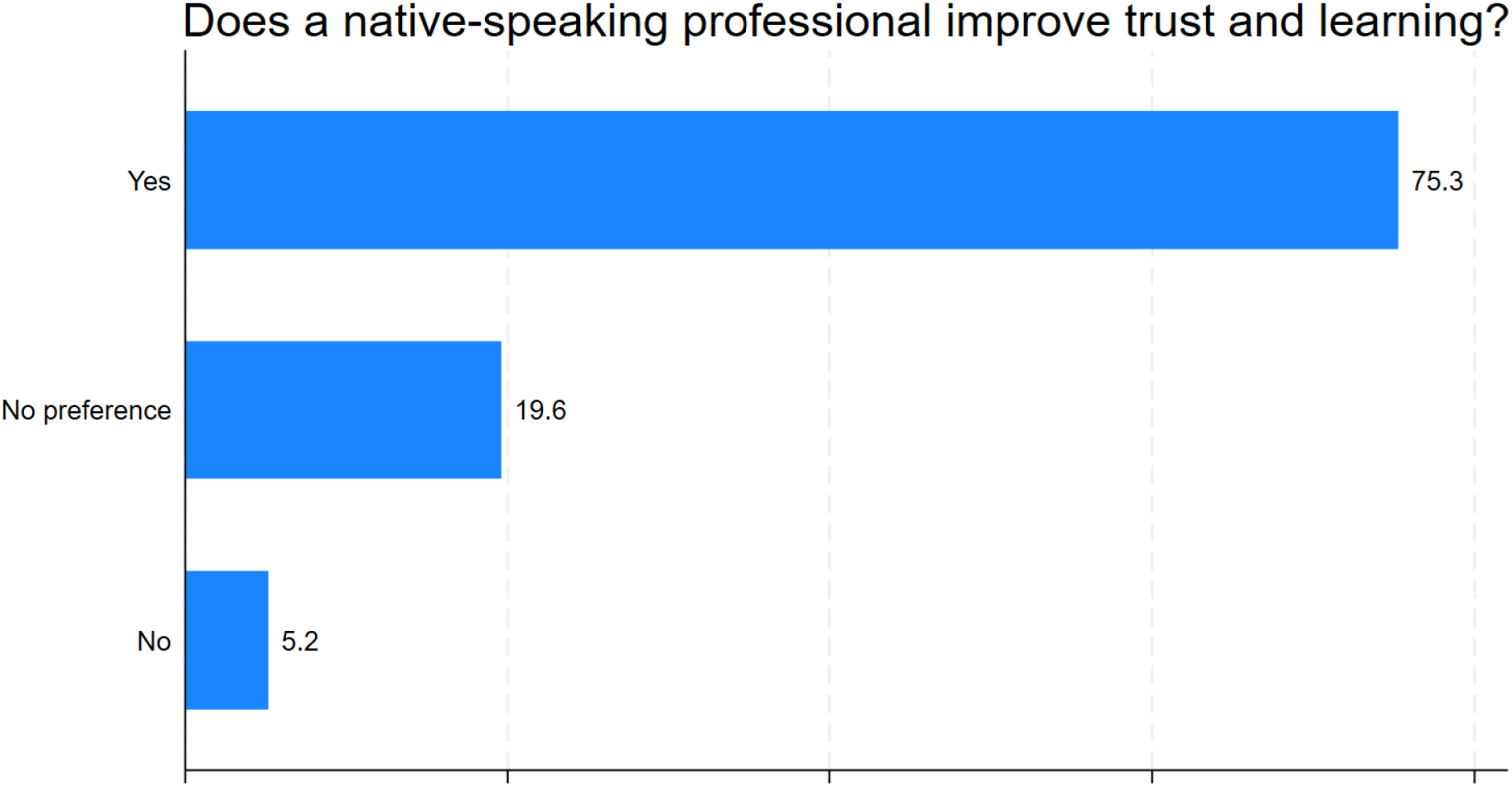
**Perceived impact of native-speaking health professionals on trust and learning**

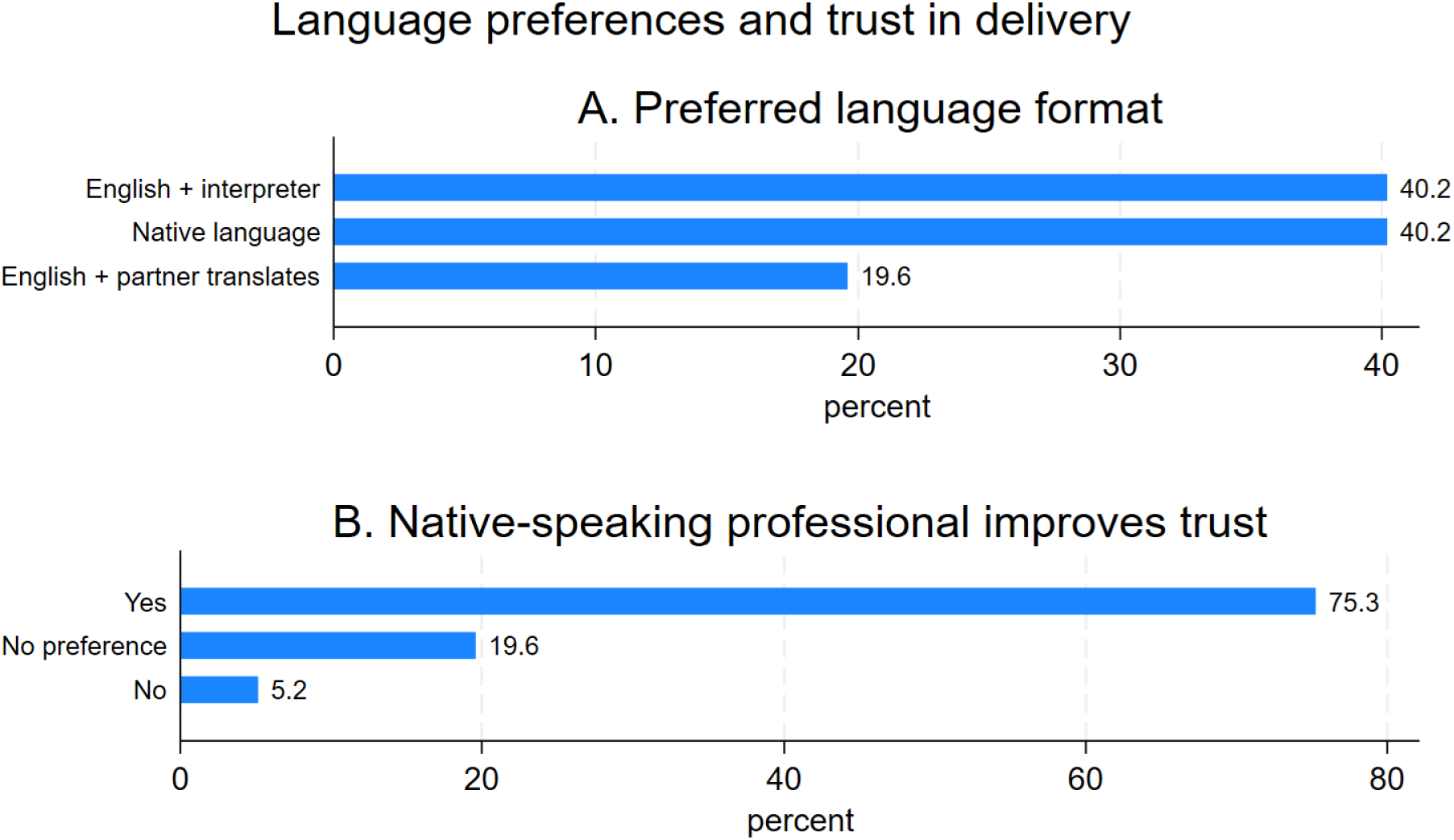
Combined Graphs figure 3 and 4: Language preferences and trust in native-speaking professionals for parent education delivery.

#### Perceived relevance of parent education content

Across all content areas assessed, the vast majority of women rated topics as important or extremely important (Table 4). Topics most frequently rated as “extremely important” included induction and augmentation of labour (45.4%), complications of labour (44.3%), and the first few hours and days after birth (45.4%). Mental health, pelvic health, and postnatal care and community support were also highly valued, with over 95% of women rating each as at least important. Few participants rated any topic as “not at all important,” indicating broad perceived relevance of parent education content.

**Table 4:** Perceived importance of parent education content across pregnancy and postnatal topics (Q34-35)

| Topic Q34 | Extremely important<br>n (%) | Very Important<br>n (%) | Important<br>n (%) | Not very important<br>n (%) | Not at all important<br>n (%) |
| --- | --- | --- | --- | --- | --- |
| Rights to maternity and health care in the UK | 33 (34.0) | 21 (21.7) | 38 (39.2) | 5 (5.2) | 0 |
| Pregnancy journey | 38 (39.2) | 33 (34.0) | 25 (25.8) | 0 | 1 (1.0) |
| When to call/access hospital | 37 (38.1) | 33 (34.0) | 27 (27.8) | 0 | 0 |
| Care meeting your own needs and wishes | 37 (38.1) | 26 (26.8) | 31 (31.9) | 0 | 3 (3.1) |
| Early labour and pain relief | 41 (42.3) | 24 (24.7) | 31 (31.9) | 1 (1.0) | 0 |
| Induction/augmentation | 44 (45.4) | 23 (23.7) | 28 (9) | 0 | 2 (2.1) |
| Complications of labour | 43 (44.3) | 27 (27.8) | 26 (26.8) | 0 | 0 |
| First few hours/days after birth | 44 (45.4) | 27 (27.8) | 26 (26.8) | 0 | 0 |
| Mental health | 41 (42.3) | 19 (19.6) | 35 (36.1) | 0 | 2 (2.1) |
| **Pelvic health** | 40 (41.2) | 18 (18.6) | 38 (39.2) | 0 | 1 (1.0) |
| **Postnatal care/community support** | 32 (32.9) | 29 (29.9) | 35 (36.1) | 0 | 1 (1.0) |

### Barriers to attendance

Reported barriers to attendance are summarised in Supplementary Table A1. Childcare was the most commonly reported barrier (42.9%), followed by transport (30.6%), timing (28.6%), and language barriers (27.1%). Digital exclusion remained relevant, with 14.4% reporting lack of reliable device or internet access and 22.7% reporting low confidence with online platforms. Multiple barriers were commonly reported.

#### Information preferences, influencers, and experience of classes

Most women expressed a preference for receiving written information in their native language (61.9%), while 29.9% preferred written information in English (Supplementary Table A2). Trusted sources of information included midwives or doctors (70.1%), printed leaflets (67.0%), social media (54.6%), and the NHS website (49.5%). Among women who attended classes, most would recommend them to others (87.6%). Many reported that they would have liked an additional support person present, most commonly a partner or husband (70.1%), followed by a female relative (18.6%).

#### Qualitative summary

Q17, Q27–28, Q31, Q33, Q35–36 were free text and some women’s responses overlapped. Thematic analysis was used to review similarities women expressed. Cultural adaptation seemed important to women regardless of their preferred language, as well as having their partner/husband and 19 women reported wishing they could bring their mother or sister to parent education classes.

## Appendix - Supplementary tables

**Table A1.** Reported barriers to attendance at NHS parent education classes (Q20-23, Q33)

| Barrier | n (%) |
| --- | --- |
| Transport (Q20) | 21 (30.6) |
| Work commitments (Q20) | 15 (21.4) |
| Childcare (Q20) | 30 (42.9) |
| Timing (Q20) | 20 (28.6) |
| Language barrier (Q20) | 19 (27.1) |
| Limited internet / device access (no-Q21) | 14 (14.4) |
| Not confident using online platforms (no -Q22) | 22 (22.7) |

\*Multiple responses permitted.

**Table A2.**
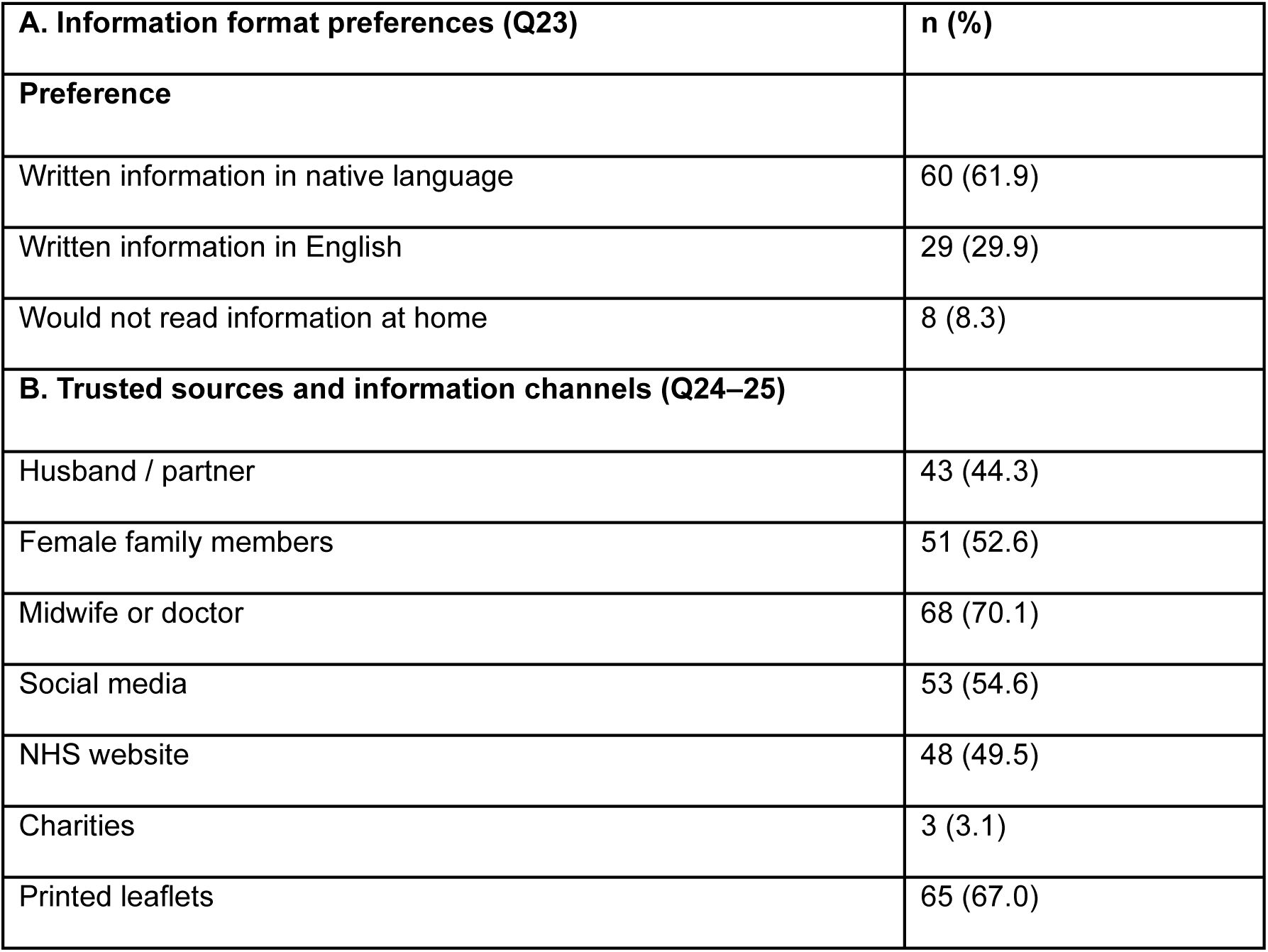
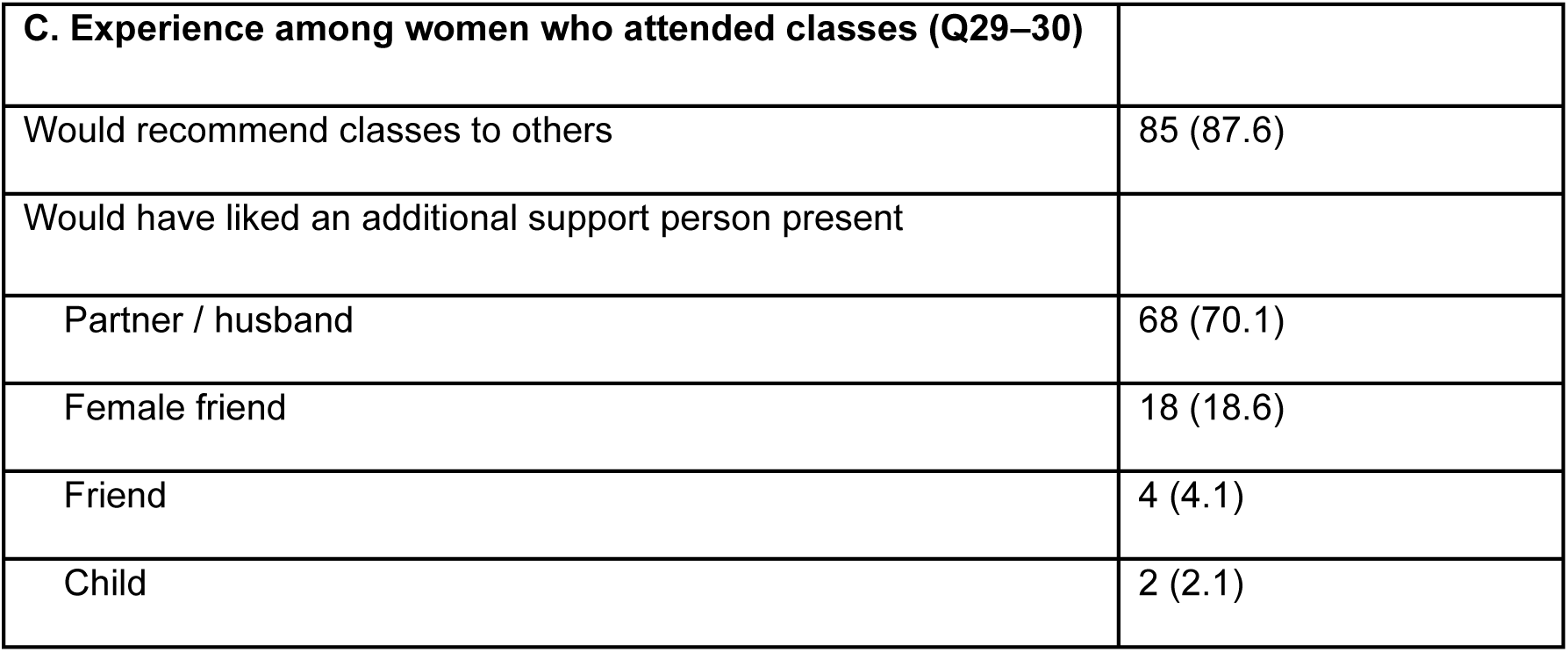
Information preferences, trusted sources, and experience of NHS parent education classes (Q23-25, Q29-30)

**Table A3.** Themes identified from free-text responses on access, relevance, and improvement of parent education services (Q17, Q27–28, Q31, Q33, Q35–36)

| **Theme** | **No. of responses** | **Example quotation** |
| --- | --- | --- |
| **Language accessibility** | n =6 | *“Language. I couldn’t feel confident to understand lesson in English.”*<br>**(Q33)** |
| **Cultural relevance** | n= 6 | *“All cultures are different. It would be very confusing to mix things as per UK norms. But beliefs should be discussed to see whether they are scientifically relevant.”* **(Q36)**<br>*“I’m a minority (Chinese) and nothing is ever properly inclusive of my culture. NHS standards for post-natal care are so low that my Chinese families think the advice given will cause long term health damage to the* |
|  |  | women by Chinese standards. Look into Chinese confinement practices and the tender care given to the emotional, mental and physical aspects to the mother during the 30 days post-natal period! I think African cultures also have similar beliefs.”<br>**(Q36)**<br>“Mention if any supplements to baby have pork or animal derived substances” **(Q36)**<br>“Maybe if they tell us what our beliefs says about pregnancy, labour and becoming a mother because a lot of the time people get confused with religion and culture” **(Q36)** |
| **Partner / family inclusion** | My mum/sister<br>n=19<br>Partner/ Husband<br>n= 70 |  |
| **Q17 Who can help midwives promote parent education classes in your community?**<br>**Other women in my community** | n=13 | “My parents consent for me to go”<br>“People who have attended the classes can help the new mothers to |
| **My Husband/partner** | n=2 | *be understand the importance of it by having a group session by calling parents who have already attended and parents to be who should be attending it.”* |
| **My family/ In Laws** | n=5 |  |
| **Religious leaders/Mosque** | n=5 |  |
| **Social Media** | n=2 |  |
| **Q27** |  |  |
| **What information did you find useful in your parent education class?** |  | *“That men can change nappies and learn to do this”* |
| **All of it was useful** | n=22 |  |
| **Early , stages & position in labour** | n=11 |  |
| **Feeding my baby and early days with a newborn** | n=7 |  |
| **Pain relief** | n=6 |  |
| **Skin to Skin** | n= 4 |  |
| **Induction** | n= 3 |  |
| **Doctors' role** | n= 2 |  |
| **Father's role** | n= 1 |  |

### Analysis

Notably, this exploratory survey is the first published patient engagement project to demonstrate that women are willing to travel to another maternity unit to access parent education in their preferred language. This highlights the importance of cross-unit partnerships and language hubs as a feasible strategy to improve equitable access to parent education for women with limited or no English proficiency. This study highlights the importance of language, cultural considerations, and accessibility in shaping engagement with NHS parent education classes among women with limited or no English proficiency.

Consistent with previous research (Turin et al., 2021; Kenyan et al., 2024), language barriers were a significant factor limiting participation. Fewer than half of participants reported access to classes with interpreter support, and nearly one in five women indicated that classes were not offered to them. Women expressed strong preferences for delivery in their native language or in English with an interpreter, and most perceived that classes led by a native-speaking midwife or female interpreter improved trust, learning, and engagement.

Cultural and family dynamics also influenced participation. Women highlighted the value of including partners, in-laws, or female relatives in parent education, reflecting earlier findings that intergenerational beliefs and household decision-making can affect use of maternity services (Goodwin et al., 2017; Nagesh et al., 2024). Many participants noted a desire for culturally adapted content that acknowledges traditional practices while providing evidence-based guidance. These findings underscore the importance of designing parent education programs that are both culturally sensitive and linguistically accessible.

Timing, format, and location were additional determinants of engagement. Women preferred earlier access to parent education, ideally before 32 weeks’ gestation, and demonstrated flexibility regarding class location, with over two-thirds willing to attend classes outside their booking unit. In-person delivery was favoured by the majority, yet high levels of digital access and confidence suggest that hybrid models could provide a practical alternative, particularly where travel or childcare pose barriers. Barriers such as childcare, transport, timing, and digital exclusion remained significant and reinforce the need for flexible, family-centered approaches.

Participants rated all content areas as relevant, with labour complications, induction, and early postnatal care being particularly important. The findings demonstrate that parent education fulfils an essential informational need. Women valued written materials in their native language, interactive teaching methods, and opportunities for peer and family support, emphasising the need for multilingual, accessible resources tailored to literacy and language proficiency.

Collectively, the findings indicate that coordinated, inclusive strategies are needed to improve equitable access. Partnerships between maternity units could standardise core content, designate language hubs, and provide culturally adapted delivery, reducing duplication and extending support to vulnerable groups. Such approaches align with national policy priorities (Ockenden Review, 2022; RCOG, 2025) and address known disparities in maternal and neonatal outcomes for migrant women with limited English proficiency.

Finally, the study underscores the value of co-production and active public involvement. Incorporating the perspectives of women, families, and Maternity & Neonatal Voice Partners (MNVPs) ensures interventions reflect the lived experiences of diverse populations.

Providing parent education in accessible, culturally sensitive, and trustworthy formats empowers women, enhances informed decision-making, and contributes to reducing health inequalities. As the UK maternity population becomes increasingly diverse, services must adapt to meet these needs while maintaining high-quality, evidence-based care.

#### Limitations

Several limitations should be considered when interpreting these findings. The sample size was relatively small (n = 97), which may limit generalisability beyond the participating London maternity networks. Although the host Trust records 49 languages among patients, the survey was translated into only 18, covering the most spoken languages. Participants could also use a language line with a healthcare professional if needed. Six translations were reviewed by native-speaking health professionals and deemed accurate, while Google Translate was used for others; the survey’s simple, clear questions minimised potential ambiguity.

Missing data is another limitation. Sociodemographic questions were optional to reduce respondent burden and protect participant comfort, resulting in missing responses, particularly for ethnicity and migration status (53%), education level (16%) and marital status, parity, and pregnancy stage (each 8%). Analyses were conducted using complete-case data for each variable, with denominators reported accordingly. Future surveys could consider making key demographic items mandatory to improve completeness.

Despite these limitations, the main descriptive findings such as language preferences, willingness to travel, and perceived relevance of parent education content were consistent and likely reflect genuine patterns. This study provides valuable insights into the needs and preferences of multilingual women using London maternity services and informs future research and service development to improve equitable access.

## Discussion

This scoping review has engaged with service users and confirmed that women will be prepared to travel to another maternity unit. This creates a unique opportunity to redesign the current fragmented landscape of antenatal education. A model based on collaboration rather than duplication could substantially improve access, efficiency, and quality. We propose exploring whether selected maternity units could act as specialist language hubs, each offering parent education in the languages with the highest local demand.

## Data Availability

All data produced in the present study are available upon reasonable request to the authors
All data produced in the present work are contained in the manuscript
All the authors have access to the data and can send you the anonymous data if requested

https://www.iolanthe.org/grants-awards/our-award-winners/2025-award-winners

